# Prevalence of Hypertension, awareness, treatment, and blood pressure control in Sierra Leone: A systematic review and meta-analysis

**DOI:** 10.1101/2024.01.05.24300711

**Authors:** Theresa Ruba Koroma, James Baligeh Walter Russell, Sallieu Kabay Samura, Joshua M Coker, Sorie Conteh, George A. Yendewa, Durodami Radcliffe Lisk

## Abstract

**Background:** In recent years, the prevalence of Hypertension (HTN) has increased in sub-Saharan countries. However, reports on the prevalence of HTN in Sierra Leone are limited. Therefore, we conducted a systematic review and meta-analysis to assess the overall estimates of the prevalence of HTN in Sierra Leone.

**Method:** A systematic search of electronic databases (PubMed, Embase, African Journal Online and Google Scholar) was conducted by three independent investigators using keywords such as “hypertension”, “prevalence”, “blood pressure”, and “Sierra Leone”. A random-effects model was used to estimate the prevalence across studies. Heterogeneity among studies was assessed using the Cochran Q test and I^2^ statistic, and publication bias was assessed using funnel plots and the Egger test.

**Results:** We pooled the data from 15 studies (n=14,707) that met the inclusion criteria. The overall prevalence of HTN in Sierra Leone was 27.64% [95% CI = 27.45, 27.82], with significant heterogeneity observed among studies (I^2^ =99.82%, p-value < 0.001). The pooled prevalence of Hypertension among males was higher than that among females (25.11% [95% CI = 24.90-25.32], I^2^ =99.79%, p-value < 0.001) and 24.11% [95% CI = 23.92,24.31, I^2^ =99.67%, p-value < 0.001]). The prevalence of HTN among urban residents (29.76%) was almost twice that among their rural counterparts (15.77%). Of those with HTN, 37.21% were aware of their illness, 12.87% received treatment, and only 29.73% had blood pressure control.

**Conclusion:** More than 1 in 4 adults aged ≥ 15 years are living with HTN in Sierra Leone, with a low awareness rate and even lower treatment and control rates. Our findings highlighted the need for robust and comprehensive hypertension prevention, treatment and control policies.

## Introduction

Hypertension is the most common cardiovascular disorder and is an independent, modifiable risk factor for global disease burden and mortality [1, 2]. Hypertension is a clinical disorder in which blood pressure (B.P.) is abnormally high following repeated measurements and is defined as systolic B.P ≥ 140 mmHg and, or diastolic B.P ≥ 90 mmHg.[3, 4] The available emerging data from several studies suggest variations in the prevalence of hypertension worldwide [5–7]. Globally, the estimated prevalence of adult hypertension in 2010 was 31.1% (1.39 billion), with a higher prevalence of 31.5% (1.04 billion) reported among adults in low- and middle-income countries (LMICs) than among those in high-income countries, with a prevalence of 28.5% (349 million) [8]. The global differences in the prevalence of hypertension could be attributed to an increased exposure to unhealthy dietary lifestyle risk factors such as high sodium consumption, low potassium intake and low physical activity, which may result from a sedentary lifestyle [9, 10]. Additionally, the ageing population and inadequate healthcare systems available in most LMICs may also contribute to the high disease burden compared with the global average [11].

By 2025, the number of hypertensive adults is expected to increase to 1.56 million, with a projected estimate of 80% of the world’s hypertensive population residing in LMICs [12]. In the last few decades, a similar trend has also been observed in sub-Saharan Africa [2]. Nevertheless, a large proportion of the population with hypertension is undiagnosed, untreated, or inadequately treated, hence contributing to the rising burden of cardiovascular disease in the region [13, 14]. The lack of awareness of hypertension among patients and non-compliance with medications, culminating in inadequate knowledge about hypertension among healthcare professionals, may contribute to poor blood pressure control [15]. A systematic analysis of hypertension revealed a higher prevalence in an urban setting in industrialized countries, with similar trends documented in developing countries [16]. Using United Nations Population Fund (UNFPA) data, Twagirumukiza et al. reported a projected increase of 68% in the prevalence of Hypertension in SSA, from 75 million (95% CI 65-93 million) in 2008 to 125.5 million (95% CI 111.0-162.9 million) by 2025 [17]. To mitigate the burden of hypertension in Africa, the Pan-African Society of Cardiology identified ten action points, and if these were to be implemented by African Ministers, there would be a 25% reduction in the burden of Hypertension by the end of 2025 [18].

Sierra Leone has been challenged by an increasing number of non-communicable diseases, and the Disability Adjusted Life Years (DALYs) in 2017 was 28.2% [19]. It was also documented that since 1990, the proportion of combined disease burden due to NCDs and injuries has risen from 22.5% to 33.9%, while the proportion of deaths has risen significantly from 28.8% to 39.6% [19]. According to the 2009 STEPS survey, approximately 35% of adults aged 25-64 years have hypertension. Nearly 20% of adults had three cardiovascular risk factors: smoking, alcohol consumption, and being overweight (World Health Organization (WHO) STEPS Survey, 2009). [20]. Currently, cardiovascular diseases account for 14% of all-cause mortality [20]. Between 1983 and 1992, approximately 7.5% of all deaths in Freetown, the capital of Sierra Leone, were due to hypertension [21]. In a retrospective study reported by Lisk in 1993, hypertension accounted for more than 65% of stroke admissions. According to a population-based study of adults in Sierra Leone, the rate of hypertension was greater than 40%. [22] A later study in an urban hospital showed a lower prevalence of 25.2% in patients older than 15 years, with an age-adjusted prevalence of 19.6%[23].

Although several studies have provided insight into the prevalence of Hypertension in Sierra Leone, no study has reviewed its trajectory over the years. Therefore, we aimed to fill this evidence gap by estimating the pooled prevalence of Hypertension at the national level by using the results reported from smaller regional studies and by investigating awareness, treatment, and blood pressure control in Sierra Leone.

## Methodology

The study protocol was registered on the International Prospective Register of Systematic Reviews (PROSPERO) with registration number CRD42023440425. This review was conducted in accordance with the Preferred Reporting Items for Systematic Reviews and Meta-Analyses (PRISMA) guidelines to ensure rigour and minimize the risk of bias [24].

### Study setting, search strategy and selection criteria

Sierra Leone is in West Africa and covers a total area of 71,740 km^2^. The country is divided into five provincial regions: the Western Area, the Northwest region, the North region, the Eastern region, and the Southern region. We systematically searched PubMed, Embase, African Journal Online (AJOL) and Google Scholar for keywords including “hypertension”, “prevalence”, “blood pressure”, and “Sierra Leone”, respectively. The reference lists of eligible studies were also searched to identify potentially eligible studies for which the electronic search may have missed. Articles from these electronic databases were screened by three independent investigators (TRK, SKS and GAY). TRK, SKS and GAY performed the study selection. Any disagreements were resolved through mutual consensus.

### Inclusion and exclusion criteria

We considered all full-text articles published in English from database inception to 31 November 2023 on hypertension prevalence, associated risk factors, management and control in Sierra Leone were considered eligible for inclusion in this systematic review. Studies conducted among individuals aged 15 years or older were also included. Multicenter studies that included Sierra Leonean participants were included if there were adequate statistical details on the data pertaining to Sierra Leone. Reports that satisfied the inclusion criteria were eligible for the initial screening. (Fig.1). We excluded all non-human studies, studies on pregnant women or children aged less than 15 years, conference abstracts, case reports, reviews, expert commentaries and studies on Sierra Leonean communities living abroad. Additionally, eligible studies that did not have the full-length article available were excluded from the quantitative synthesis or meta-analysis.

**Fig 1:**
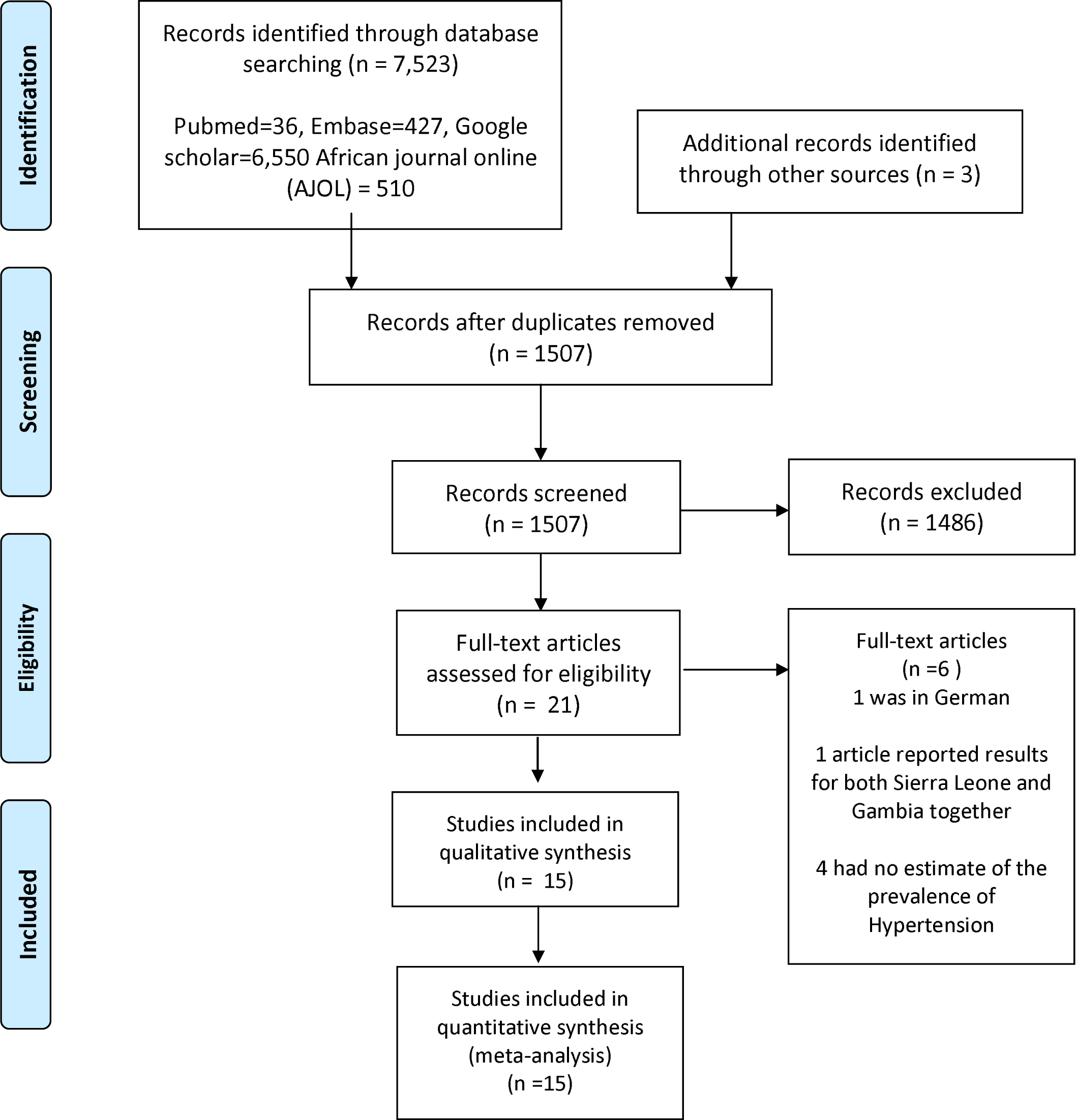
Flow diagram for studies included in the meta-analysis

### Outcome of the analysis

The primary outcome of this study was the pooled prevalence of hypertension in the general (community-based) or different subsets of the population. The secondary outcome is to assess the prevalence of awareness, treatment, and control of hypertension.

### Data extraction and quality assessment

Using Microsoft Excel, relevant data were retrieved from selected manuscripts independently by TRK, SKS, and GAY. Any discrepancies between the three reviewers were settled through discussion. For each study, the following information was extracted: authors’ names, publication date, sample size, setting, geographic location, diagnostic criteria, population studied and the overall prevalence of Hypertension. Furthermore, we performed statistical analysis of the observed differences in the prevalence of Hypertension based on sex and mean systolic and diastolic blood pressure among hypertensive patients if reported; other cardiovascular risk factors reported; and definitions for rural and urban locations as well as information for the assessment of the risk of bias. We contacted authors of studies with incomplete information online and requested manuscripts for data extraction.

In addition, TRK, SKS and GAY assessed all included studies using the Newcastle□Ottawa Scale (NOS) under the three domains of selection, comparability, and exposure [1]. The NOS contains nine items with a total maximum possible quality score of 9. We rated studies with a quality score of 9-8 as very high quality, a quality score of 7-6 as high quality, a quality score of 5-4 as moderate quality, and a quality score of 3-0 as low quality[25].

### Statistical Analysis

We used STATA version 17 for statistical analysis. Standard errors for study-specific prevalence were calculated from the point estimate and appropriate denominators, assuming a binomial distribution. Using a random-effects meta-analysis, we pooled the study-specific estimates to achieve an overall summary estimate of the prevalence across studies. Heterogeneity among studies was assessed using the Cochran Q test and I^2^ statistic [26]. The I^2^ statistic estimates the percentage of total variation across studies that is due to true between-study differences rather than chance. We explored sources of heterogeneity by comparing hypertension prevalence between subgroups defined by study-level characteristics. We assessed the presence of publication bias using funnel plots and the Egger test and by comparing the pooled prevalence between larger and smaller studies [27].

## Results

### Study selection

Our search yielded 7,523 articles from PubMed (36), Embase (427), Google Scholar (6,550) and AJOL (510). Three additional studies were identified from other sources. After excluding 6016 duplicate articles, we screened 1507 titles and abstracts for relevance, 21 of which were retrieved for full-text screening. Of the 21 studies screened, six were excluded because they did not meet the inclusion criteria of our study. One study was in German, another reported results for both Sierra Leone and Gambia together, and 4 had no estimate of the prevalence of hypertension. A total of 15 studies were ultimately included in this systematic review and meta-analysis. (Figure 1)

### General characteristics of the included studies

The characteristics of the 15 studies [23, 28–41] included in this review are summarized in Table 1. A total of 14,707 participants aged ≥ 15 years were included in all fifteen studies, with sample sizes ranging from 87 to 3944. Five retrospective, one prospective and seven cross-sectional surveys were included in this meta-analysis. Two of the studies did not specify their study design. Hypertension prevalence was reported as a primary outcome in all fifteen studies, whereas five reported on awareness, 2 on treatment and 4 on hypertension control. The NOS quality assessment for the included studies is presented in Table 2. Two studies (13%) were rated as very high, 10 (67%) were rated as high and 3 (20%) were rated as moderate quality. Publication bias was evaluated graphically using a funnel plot (2) and the meta-regression Egger test to determine the potential source of heterogeneity (Z= 1.89, pvalue = 0.0584).

**Table 1:**
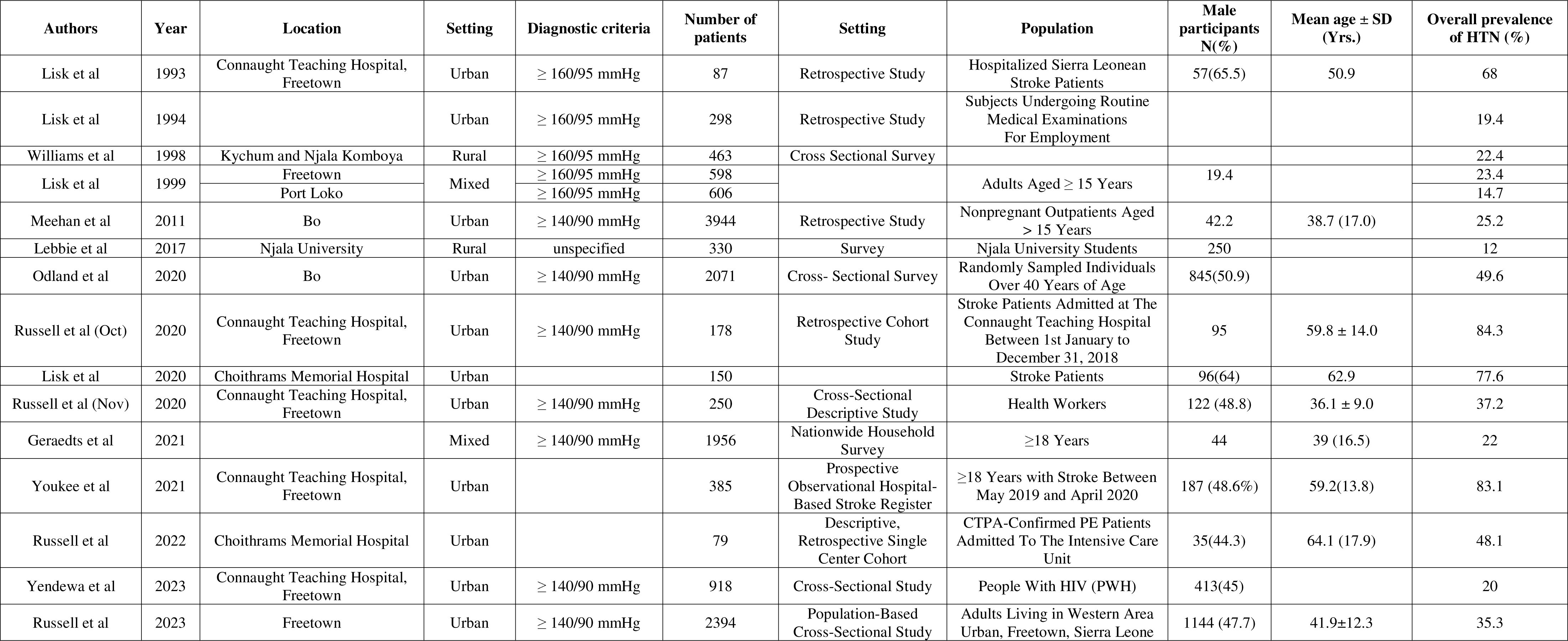
Study characteristics of patients with Hypertension.

| Authors | Year | Location | Setting | Diagnostic criteria | Number of patients | Setting | Population | Male participants N(%) | Mean age ± SD (Yrs.) | Overall prevalence of HTN (%) |
| --- | --- | --- | --- | --- | --- | --- | --- | --- | --- | --- |
| Lisk et al | 1993 | Connaught Teaching Hospital, Freetown | Urban | ≥ 160/95 mmHg | 87 | Retrospective Study | Hospitalized Sierra Leonean Stroke Patients | 57(65.5) | 50.9 | 68 |
| Lisk et al | 1994 |  | Urban | ≥ 160/95 mmHg | 298 | Retrospective Study | Subjects Undergoing Routine Medical Examinations For Employment |  |  | 19.4 |
| Williams et al | 1998 | Kychum and Njala Komboya | Rural | ≥ 160/95 mmHg | 463 | Cross Sectional Survey |  |  |  | 22.4 |
| Lisk et al | 1999 | Freetown | Mixed | ≥ 160/95 mmHg | 598 |  | Adults Aged ≥ 15 Years | 19.4 |  | 23.4 |
|  |  | Port Loko |  | ≥ 160/95 mmHg | 606 |  |  |  |  | 14.7 |
| Meehan et al | 2011 | Bo | Urban | ≥ 140/90 mmHg | 3944 | Retrospective Study | Nonpregnant Outpatients Aged > 15 Years | 42.2 | 38.7 (17.0) | 25.2 |
| Lebbie et al | 2017 | Njala University | Rural | unspecified | 330 | Survey | Njala University Students | 250 |  | 12 |
| Odland et al | 2020 | Bo | Urban | ≥ 140/90 mmHg | 2071 | Cross- Sectional Survey | Randomly Sampled Individuals Over 40 Years of Age | 845(50.9) |  | 49.6 |
| Russell et al (Oct) | 2020 | Connaught Teaching Hospital, Freetown | Urban | ≥ 140/90 mmHg | 178 | Retrospective Cohort Study | Stroke Patients Admitted at The Connaught Teaching Hospital Between 1st January to December 31, 2018 | 95 | 59.8 ± 14.0 | 84.3 |
| Lisk et al | 2020 | Choithrams Memorial Hospital | Urban |  | 150 |  | Stroke Patients | 96(64) | 62.9 | 77.6 |
| Russell et al (Nov) | 2020 | Connaught Teaching Hospital, Freetown | Urban | ≥ 140/90 mmHg | 250 | Cross-Sectional Descriptive Study | Health Workers | 122 (48.8) | 36.1 ± 9.0 | 37.2 |
| Geraedts et al | 2021 |  | Mixed | ≥ 140/90 mmHg | 1956 | Nationwide Household Survey | ≥18 Years | 44 | 39 (16.5) | 22 |
| Youkee et al | 2021 | Connaught Teaching Hospital, Freetown | Urban |  | 385 | Prospective Observational Hospital-Based Stroke Register | ≥18 Years with Stroke Between May 2019 and April 2020 | 187 (48.6%) | 59.2(13.8) | 83.1 |
| Russell et al | 2022 | Choithrams Memorial Hospital | Urban |  | 79 | Descriptive, Retrospective Single Center Cohort | CTPA-Confirmed PE Patients Admitted To The Intensive Care Unit | 35(44.3) | 64.1 (17.9) | 48.1 |
| Yendewa et al | 2023 | Connaught Teaching Hospital, Freetown | Urban | ≥ 140/90 mmHg | 918 | Cross-Sectional Study | People With HIV (PWH) | 413(45) |  | 20 |
| Russell et al | 2023 | Freetown | Urban | ≥ 140/90 mmHg | 2394 | Population-Based Cross-Sectional Study | Adults Living in Western Area Urban, Freetown, Sierra Leone | 1144 (47.7) | 41.9±12.3 | 35.3 |

**Table 2:**
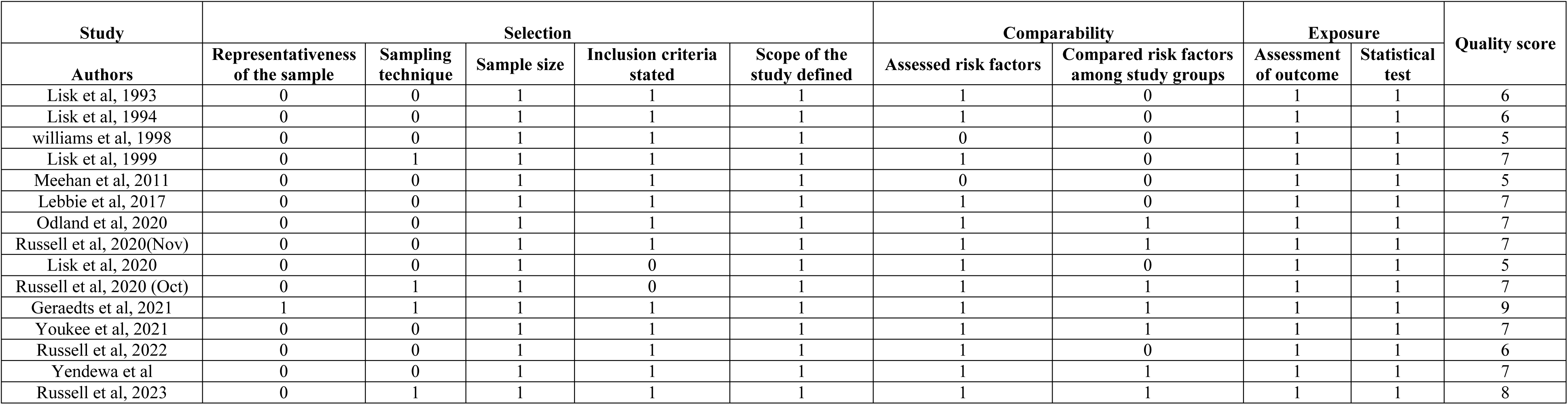
Quality assessment of included studies using the Newcastle-Ottawa Scale (NOS)

Six studies defined hypertension as a blood pressure (B.P.) ≥ 140/90 mmHg, four studies used the old WHO definition of B.P.≥ 160/95 mmHg, and five studies did not specify the B.P. cut-off. Four studies(n=1965) were from the 1990s, and eleven studies (n= 12742) were published after 2010. Two of the included studies were conducted in an urbanrural mixed setting, two other studies reported data from only rural areas, and the remaining eleven were from urban settings. The mean age reported among studies ranged from 36 to 64 years.

There was wide variation in the protocols used for B.P. measurement regarding the intervals between B.P. measurements, frequency, body posture, part of the body on which the B.P. was taken, and the type of B.P. device used. The most used device for measuring B.P. was an Omron digital automatic blood pressure machine, followed by a mercury sphygmomanometer. Bp was measured at least once in all the studies, with the preferred site for B.P. measurement being the upper arm with the patient seated upright.

### Crude prevalence of Hypertension in Sierra Leone

The highest prevalence of hypertension from individual study estimates was recorded in Freetown among stroke patients admitted to the Connaught Teaching Hospital between 1st January and December 31, 2018 at 84.3% in 2020[35]. The lowest estimated prevalence was recorded at Njala University, among which 12% were university students in 2017 [31].

### Pooled prevalence of hypertension in Sierra Leone

Using a random effects model, the overall prevalence of Hypertension in Sierra Leone was 27.64% [95% CI = 27.45, 27.82] with significant heterogeneity observed among studies I^2^ =99.82%, pvalue < 0.001 (Fig 2). The highest weight among the studies included was observed in the studies conducted by Meehan et al. [23], followed by Geraedts et al. [34] and Russell et al[35].

**Fig. 2:**
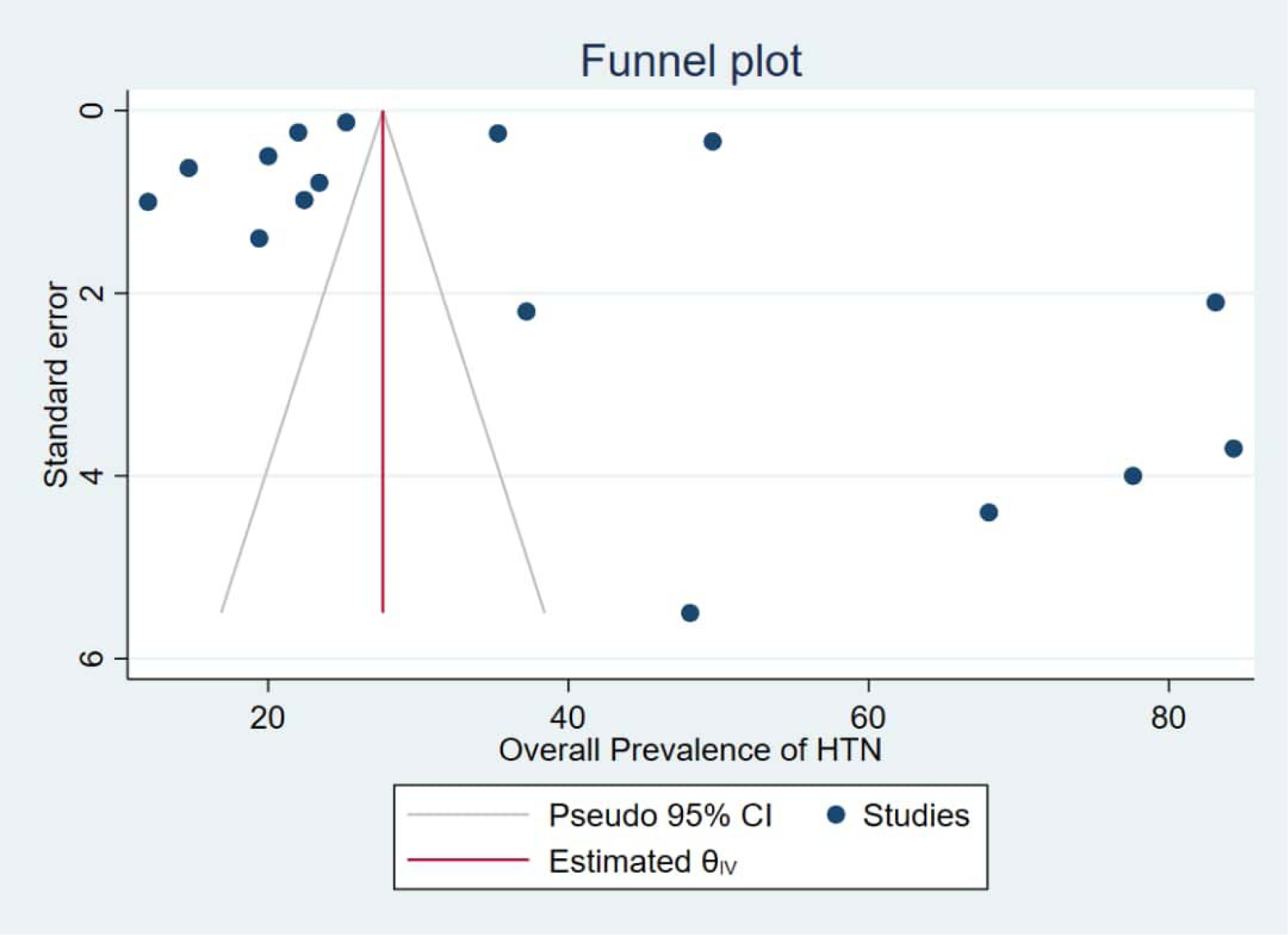
Funnel plot of prevalence of hypertension versus standard error

The pooled prevalence of hypertension among males in the five studies was 25.11% (95% CI = 24.90-25.32), with significant heterogeneity observed among studies I^2^ =99.79%, pvalue < 0.001, while that of females was 24.11% (95% CI = 23.92,24.31) with significant heterogeneity observed among studies I^2^ =99.67%, pvalue < 0.001 (Fig. 3). In general, Hypertension was more prevalent in males than in females.

**Fig. 3:**
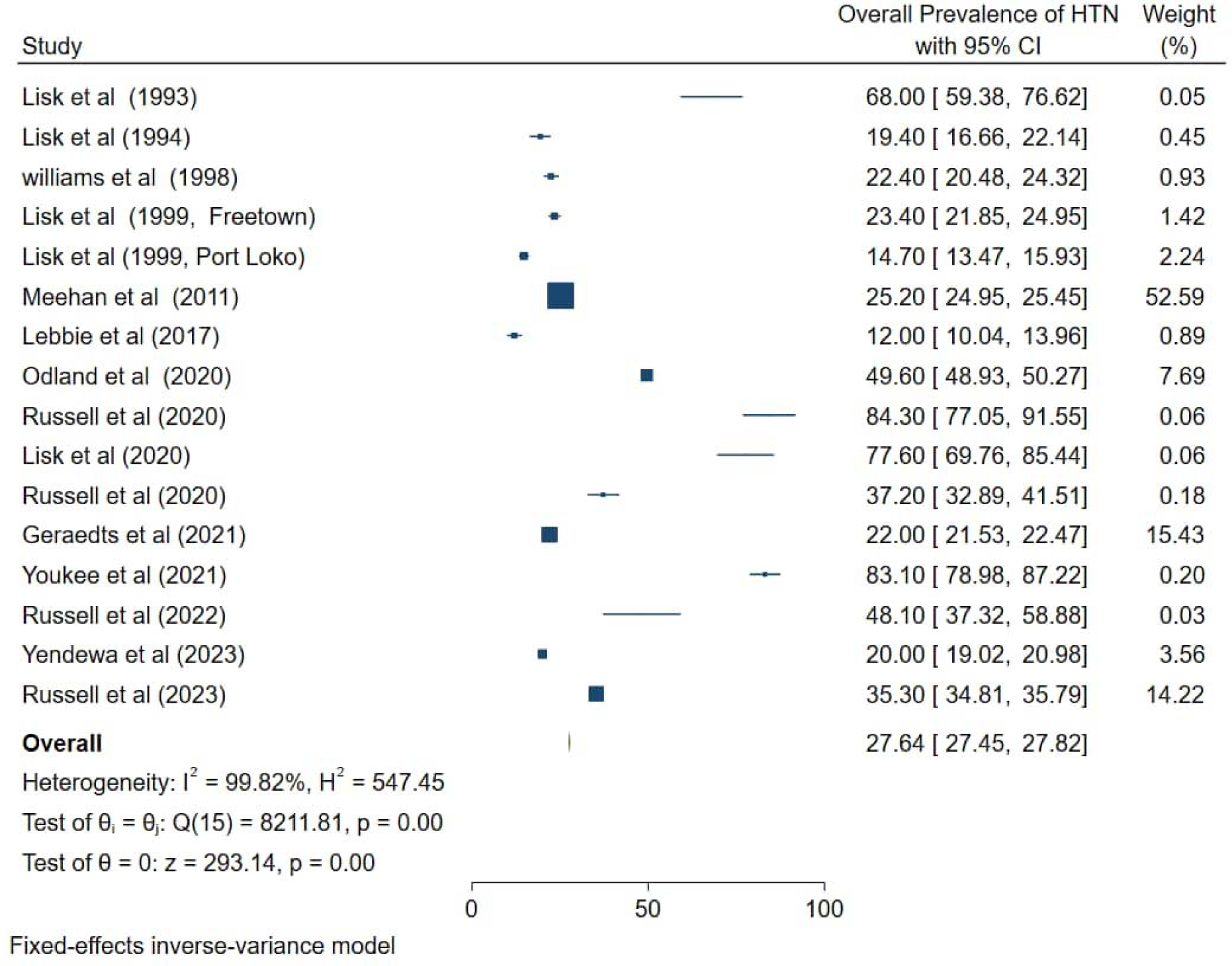
Pooled prevalence of Hypertension in Sierra Leone

The pooled prevalence of hypertension among urban residents was greater than that among their rural counterparts. We estimated prevalence rates of 29.76% (95% CI = 29.55,29.96 I^2^ = 99.85 pvalue<0.001) in the urban setting and 15.77% (95% CI = 14.88,16.67, I^2^ = 96.79, p-value<0.001) in rural areas.

### Awareness, treatment, and control of Hypertension

The pooled hypertension awareness rate was 37.21% (95% CI = 22.20. 52.22) with a significant heterogeneity (I^2^ = 99.28%, pvalue < 0.001. A total of 12.87% (95% CI = 9.25,16.50) of the patients were taking antihypertensive medications (1^2^= 89.41%, pvalue < 0.001 and 29.73% (95% CI = 880. 50.661) of those on treatment had controlled B.P. (Fig. 4)

**Fig 4:**
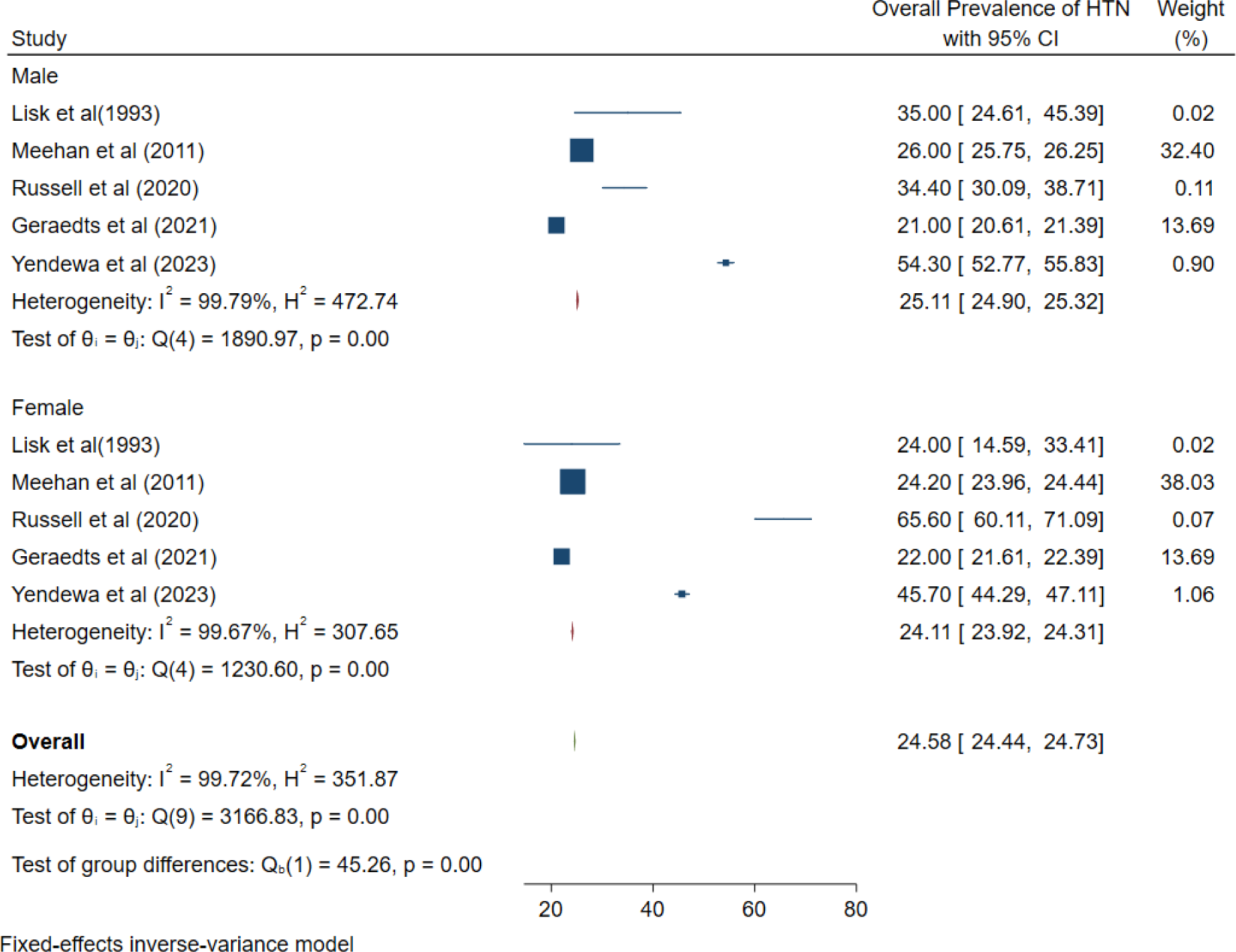
Pooled prevalence of hypertension amongst males and females in Sierra Leone

**Fig 5:**
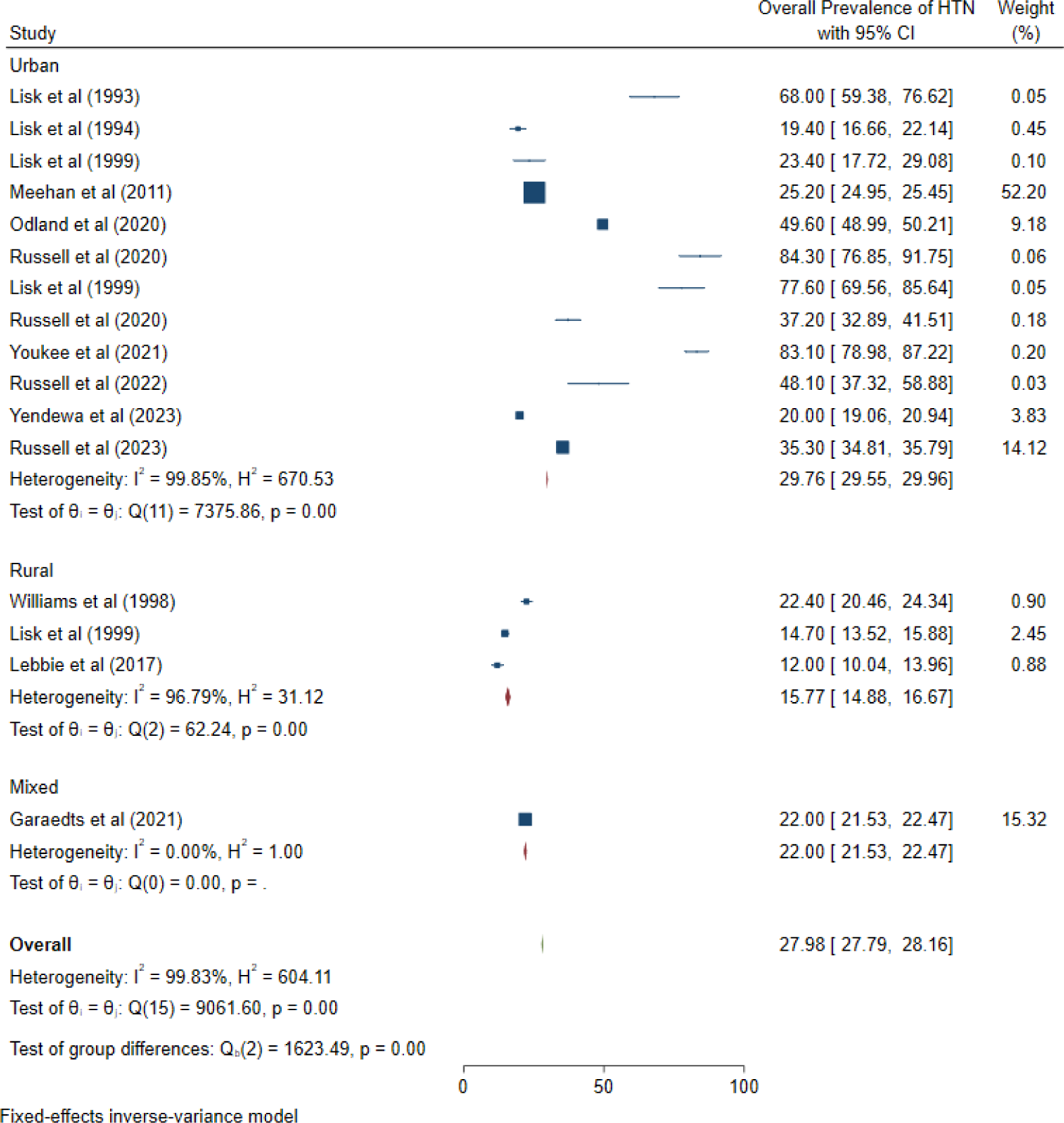
Pooled prevalence of hypertension between urban and rural areas in Sierra Leone

**Fig 6:**
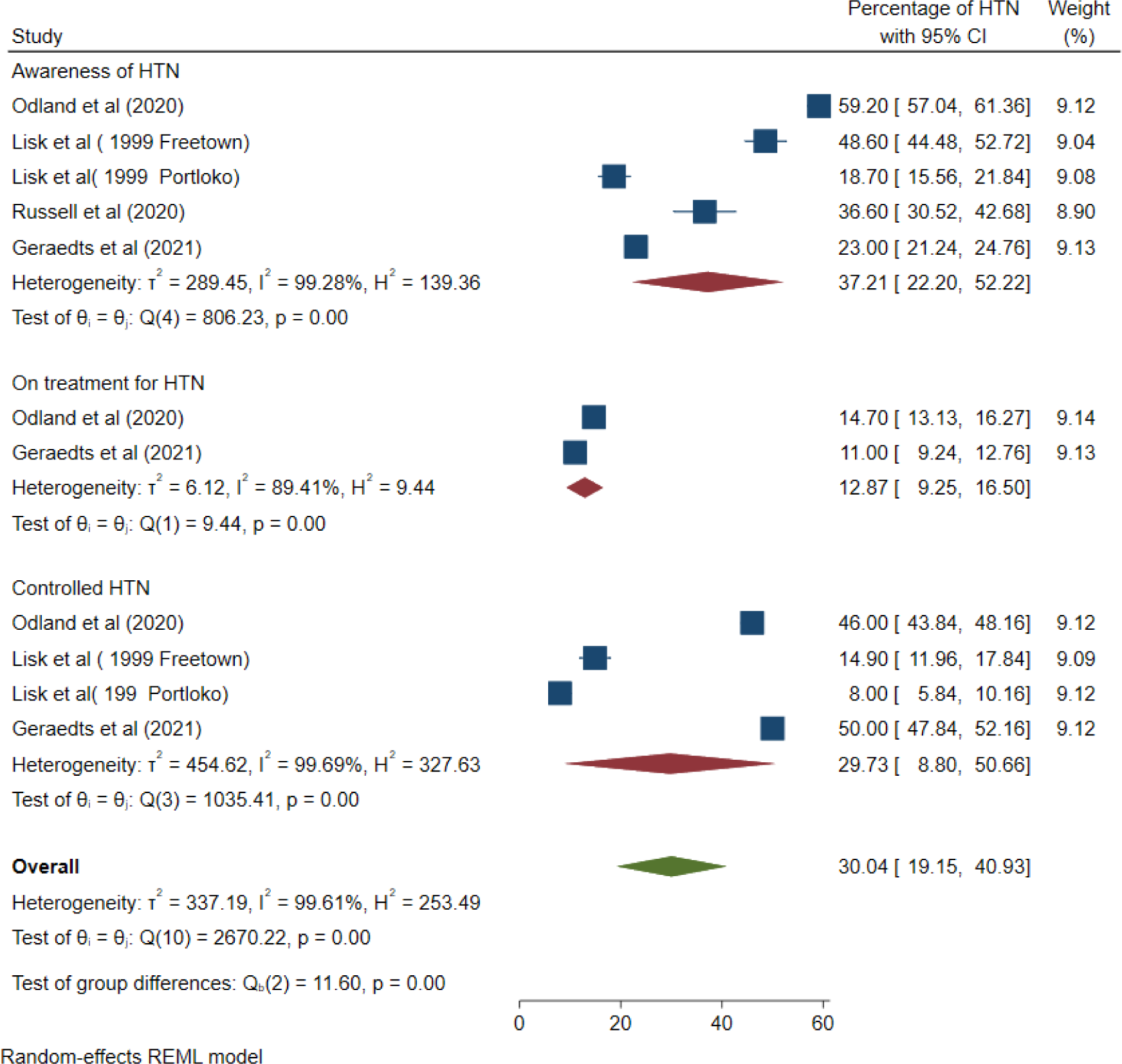
Pooled prevalence of hypertension awareness, treatment, and control amongst people with Hypertension in Sierra Leone.

## Discussion

The increase in the prevalence of cardiometabolic risk factors such as hypertension has increased the burden of cardiovascular diseases worldwide. In this study, we systematically reviewed the available literature on the of hypertension in Sierra Leone to present up-to-date information on this disease condition. We not only meta-analysed the prevalence of hypertension among the fifteen studies included but also examined the disparities in the prevalence of hypertension between rural and urban areas in Sierra Leone, as well as sex differences. The studies included were conducted in urban and rural settings involving young and elderly people. To the best of our knowledge, this is the first comprehensive review and meta-analysis on the prevalence, awareness, treatment, and control of hypertension in Sierra Leone.

Our study demonstrated a pooled prevalence of hypertension of 27.6% in Sierra Leone, with a higher prevalence in males than females. Our finding is comparable to most of the studies conducted in the general population but significantly lower than the findings of studies conducted in hospital settings[42]. Notably, the prevalence of hypertension was higher in the older population [32] and among patients with certain medical conditions (for example, stroke) [42], with the lowest prevalence rate reported in young university students (12%)[31]. To date, there has not been any meta-analysis on the prevalence of hypertension in Sierra Leone to compare our findings. However, when compared with systemic reviews in the subregion, Ogah et al[43]. estimated a prevalence of 22.5% in Nigeria for the period 2000– 2009. Adeyole et al. reported an age-adjusted hypertension prevalence of 32.5% in 2020, an increase of more than 540% from 4 million to 28 million patients between 1995 and 2020 [44]. The prevalence reported in our study is consistent with that estimated for Ghana (27%)[45] but higher than the prevalence of hypertension in Ethiopia reported by Sofonyas et al. (22%)[46]. These differences might be due to the study settings, diagnostic criteria for hypertension, the age of participants in the studies, and the length of the study.

The pooled estimate of hypertension from the random effects model was higher in males than females (25% versus 24%). The observed differences in hypertension prevalence between genders may be attributed to the protective effect of pre-menopausal female hormones, coupled with the notion that men are more susceptible to the behavioural risk factors associated with hypertension. Similar findings were reported in studies conducted in Ethiopia[46], Ghana[47] and Nigeria[48]. In contrast, a reverse trend was reported in a study conducted in Uganda [49], with Sani et al. reporting no significant differences between sexes[50].

The observed prevalence of Hypertension (30%) in the urban setting was higher than the 16% documented in rural settings. This finding is comparable to the results of several studies performed in SSA. In a recent meta-analysis on hypertension prevalence in Nigeria, the authors reported a prevalence of 31% among urban dwellers versus 26% among their rural counterparts [50].

Likely contributing factors to the variation in the prevalence of hypertension among urban and rural areas can be attributed to an increase in urbanization, differences in socioeconomic conditions, low physical activity, unhealthy diet[51], an aging population, and the lack of effective nationwide preventive measures. Easy access to healthcare facilities in urban areas may also contribute to the higher prevalence of pregnancy in these areas than in rural areas [52]. Another study on hypertension in Africa reported a significantly in urban settings than in rural settings in 1990 but no difference between 2000 and 2010, which was a result of possible reverse ruralurban migration[14]. Contrary to our findings, Noubiap et al. reported a higher prevalence of hypertension among children and adolescents in rural areas than in urban areas [53].

Most notable from our study is the low rate of awareness of hypertension. Only 37.21% were aware of their status before enrollment in the studies, 12.87% were receiving treatment, and 29.73% had controlled B.P. The low awareness rate observed in our study is in keeping with findings from studies performed in SSA countries. A recent national survey in Kenya showed that only 15% of individuals with hypertension were aware of their elevated blood pressure, and only 27% were receiving treatment [54]. Nonetheless, the proportion of hypertension awareness was greater among older respondents, obese individuals, and female respondents.

In contrast to our findings, an upwards trend of up to 80% in the rate of awareness has been reported for the NCD risk factor collaboration (NCD-RisC) over the past few decades in 12 high-income countries.[55] Community outreach and screening for hypertension might aid greatly in educating the masses on hypertension. In addition, universal healthcare coverage or health insurance might be essential for improving access to care.

Approximately a quarter of those on treatment were observed to have successful B.P. control. This can be because of inadequate patient understanding of hypertension, noncompliance/poor adherence to treatment, cost of treatment, and lack of structured follow-up programs for these patients. Recently, many countries have endorsed identifying and controlling HTN as a priority and the availability of affordable and effective antihypertensive medications. However, the control of B.P. still poses a challenge among those diagnosed with this condition. A study on early experiences and lessons learned from an NCD clinic in rural Sierra Leone reported an increase in the percentage of patients with controlled B.P. from approximately 7% to 31% over an average treatment duration of 35 weeks[56]. This finding indicates that access to care and a structured follow-up program can increase the rate of compliance with treatment and, hence, increase the rate of B.P. control.

### Limitations of the study

Our study contributed to an updated prevalence of hypertension in Sierra Leone. However, this study has several shortcomings. First, there were only few population-based studies of hypertension in Sierra Leone; hence, both retrospective and survey studies were included in our meta-analysis. Additionally, there were variations in the B.P cut-off for hypertension among studies, with earlier studies using a B.P. cut-off of ≥ 160/95 mmHg, which might have led to an underestimation of the true prevalence of this disease. In addition, awareness, treatment and control meta-analyses were performed for only a subset of the studies. Therefore, the findings may not be fully representative of the whole population. Finally, significant heterogeneity was observed across the included studies, possibly because of differences in study sample size, study population, and study protocols. Hence, we performed an Egger meta-regression test to confirm the validity of the results of this meta-analysis.

## Conclusion

According to the findings of this systematic review, hypertension is a major health issue in Sierra Leone that requires immediate attention from public health authorities. The review provides the first country-specific estimates of hypertension in Sierra Leone and highlights the difference in prevalence between urban and rural areas. The pooled prevalence of hypertension in Sierra Leone was 27.6%, with low awareness and control of blood pressure rates. This review also reported higher rates of hypertension in male patients and urban dwellers. These findings are significant for future research and can help inform appropriate public response and educational campaigns. Furthermore, we recommend implementing a well-designed, nationwide, population-based surveillance on hypertension to provide more precise and representative estimates of its prevalence.

## Data Availability

All data produced in the present study are available upon reasonable request to the authors.

## Acknowledgement

None to declare

## Ethical approval and consent to participate

Not applicable

## Author contributions

TRK and JBWR conceptualized and designed the study. TRK, SKS and GAY conducted the literature search, extracted the data, and analysed and interpreted the data. TRK wrote the first draft. SKS, TRK, JBWR, JMC, and DRL contributed to the final draft and edited the manuscript for important intellectual content. All the authors approved the manuscript for submission.

